# Climate change and the rising incidence of dengue in Argentina

**DOI:** 10.1101/2022.06.03.22275954

**Authors:** MS López, A Gómez, G Müller, E Walker, MA Robert, EL Estallo

## Abstract

**BACKGROUND:** One of the most important consequences of climate change is the increased spread of dengue from tropical and subtropical regions to temperate areas around the world. Climate variables such as temperature and precipitation influence the dengue vector’s biology, physiology, abundance, and life cycle. Thus, an analysis is needed of climate changes and their possible links with the growing occurrence of epidemics recorded in recent decades.

**OBJECTIVES:** To assess the increasing incidence of dengue driven by climate change at the southern limits of dengue virus transmission in South America.

**METHODS:** We analyze the evolution of climate, epidemiological, and biological variables, comparing a period without the presence of dengue cases to a more recent period with the occurrence of cases and, consequently, of important epidemics. Temperature and precipitation are the climate variables evaluated, the total number of cases and incidence of dengue are the epidemiological variables, and finally, the optimal temperature ranges for transmission of the dengue vector is the biological variable.

**RESULTS:** The presence of dengue cases and epidemic outbreaks are observed to be consistent with positive temperature trends and anomalies. Dengue cases do not seem to be associated with precipitation trends and anomalies. The number of days with optimal temperatures for dengue transmission increased from the period without dengue cases to the period with the presence of dengue cases. However, the number of months with optimal transmission temperatures was the same in both periods.

**CONCLUSIONS:** The higher incidence of dengue virus (DENV) and its expansion to different regions of Argentina seem to be associated with temperature increases in the country during the past decades. The active surveillance of both the vector and associated arboviruses will make it possible to assess and predict the occurrence of epidemics, based on the accelerated changes in climate. Such surveillance should go hand in hand with efforts to improve the understanding of the mechanisms driving the geographic expansion of dengue and other arboviruses beyond the current limits.

---

Climate can affect the transmission dynamics, geographic spread, and re-emergence of vector-borne diseases (Rocklöv and Dubrow 2020). In the past few decades, many mosquito-borne diseases have expanded their distributions from tropical and subtropical regions to temperate areas around the world (La Ruche et al. 2010; Tomasello et al. 2013; Rey 2014; Robert et al. 2019; Robert et al. 2020; López et al. 2021). The factors that contribute to the global expansion and intensification of dengue virus (DENV) and other arboviruses include global-scale travel, rapid and unplanned urbanization, and changes in climate leading to increased temperatures and erratic precipitation patterns (Gubler 2002, 2011; Hii et al. 2009; Butterworth et al. 2017; Huber et al. 2018). Dengue fever is considered one of the most important emerging and re-emerging arboviral diseases at present (Gubler 2020; WHO 2020). In addition, severe effects on health due to increasing dengue epidemics are predicted for the next several decades as a consequence of projected climate changes (IPCC 2022). The Americas is one of the most severely affected regions, with the southern limit of dengue virus (DENV) transmission located in Argentina, South America (WHO 2020).

The first records of DENV transmission in Argentina date back to 1916 and include an outbreak in the northeast of the country. Dengue transmission was reconfirmed some 70 years later at the end of the 1990s when an epidemic occurred in the northern subtropical provinces (Avilés et al. 1999; Brathwaite et al. 2012). Since then, autochthonous DENV transmission has been reported in the northernmost provinces almost every year (Vezzani and Carbajo 2008). In 2009, autochthonous DENV transmission was detected for the first time in temperate central Argentina, and since then, dengue incidence has increased considerably in most of the country’s provinces (Robert et al. 2019; López et al. 2021). The expansion of dengue beyond tropical and subtropical latitudes where dengue is endemic confirms the haste in addressing this problem (Wilder-Smith and Gubler 2008; Ferreira 2012; Murray et al. 2013; Messina et al. 2014). Large-scale global dengue epidemics are associated mainly with the presence of Aedes aegypti mosquitoes, which are also responsible for transmitting other emerging and re-emerging arboviruses such as yellow fever, Zika, and chikungunya (Lambrechts et al. 2010; Patterson et al. 2016).

In Argentina, the geographical distribution of Ae. aegypti is expanding. Records show that in 1986 the vector was found in only two provinces in the north of the country above 28° S latitude (Díaz-Nieto et al. 2013). Currently, there are records of the species up to 40° S latitude (Rubio et al. 2020) in 20 of the country’s 24 provinces. Climate conditions, especially temperature, influence the global spread of the vector, the species’ life history characteristics, and the acceleration of the virus transmission capacity (Boggs et al. 2012; Bhatt et al. 2013; Carrington et al. 2013; Kraemer et al. 2015; Caldwell et al. 2021). Additionally, the extrinsic incubation period (the time between ingestion of the pathogen by the vector and the vector becoming infectious) for DENV is inversely associated with ambient temperature (Rocklöv and Dubrow 2020). According to the Sixth Assessment Report of the Intergovernmental Panel on Climate Change (IPCC 2021), each of the past four decades has been successively warmer than any decade since 1850, and global surface temperature in the first two decades of the 21st century (2001–2020) was 0.99 °C higher than in 1850–1900. Global surface temperatures will continue to rise until at least the mid-century unless deep reductions in CO_2_ and other greenhouse gas emissions occur in the coming decades (IPCC 2021). In addition, heavy precipitation and associated flooding events are projected to become more intense and frequent in some regions of South America (e.g., Lovino et al. 2018a, 2018b). These projected changes in climate conditions are predicted to affect the distribution and competence of Ae. aegypti and other vectors and have a potentially significant impact on the epidemiology of dengue (and other vector-borne diseases) globally (Rocklöv and Tozan 2019). The impact of climate change on the incidence, transmission-season duration, and spread of vector-borne diseases represents a major threat (Rocklöv and Dubrow 2020).

As global temperatures continue to rise, the concerns are that the mosquito and virus will spread to higher latitudes and DENV incidence will increase (Rocklöv and Dubrow 2020). The intensification of dengue epidemics over the past decade in regions at the southern limit of distribution of Ae. aegyti is indicative of increasing dengue emergence, and it is extremely necessary to better understand the drivers that contribute to its increase. This work aims to analyze the evolution of the incidence of DENV in Argentina from its re-introduction in 1998 until the most recent and largest epidemic in 2020, and its relation to climate change. We described the trend and anomalies of temperature and precipitation across the country. Finally, we analyzed the number of months and days with suitable temperature conditions for DENV transmission in representative cities.

## Methods

### Health data

The Argentine Ministry of Health (MoH) groups 24 provinces or federal states (https://www.argentina.gob.ar/pais/provincias) into five regions (Figure 1c). Confirmed and probable autochthonous cases of dengue in all regions in the period 1998–2020 were compiled from the MoH’s periodical National Epidemiological Bulletins (https://bancos.salud.gob.ar/bancos/materiales-para-equipos-de-salud/soporte/boletines-epidemiologicos). In those reports, we identified key data, including the cumulative number of probable dengue cases (i.e., with at least one positive laboratory diagnosis) as well as confirmed cases (with two positive laboratory tests), and autochthonous cases (i.e., locally transmitted) per year and province. This study does not include imported cases (i.e., illness in individuals with a travel history to regions with arbovirus activity), or suspected and unconfirmed cases (suspected cases have infection symptoms but lack laboratory diagnosis). The provinces with registered autochthonous DENV cases were Buenos Aires, Autonomous City of Buenos Aires, Catamarca, Chaco, Córdoba, Corrientes, Entre Ríos, Formosa, Jujuy, La Pampa, La Rioja, Mendoza, Misiones, Neuquén, Salta, San Juan, San Luis, Santa Fe, Santiago del Estero, and Tucumán.

**Figure 1.**
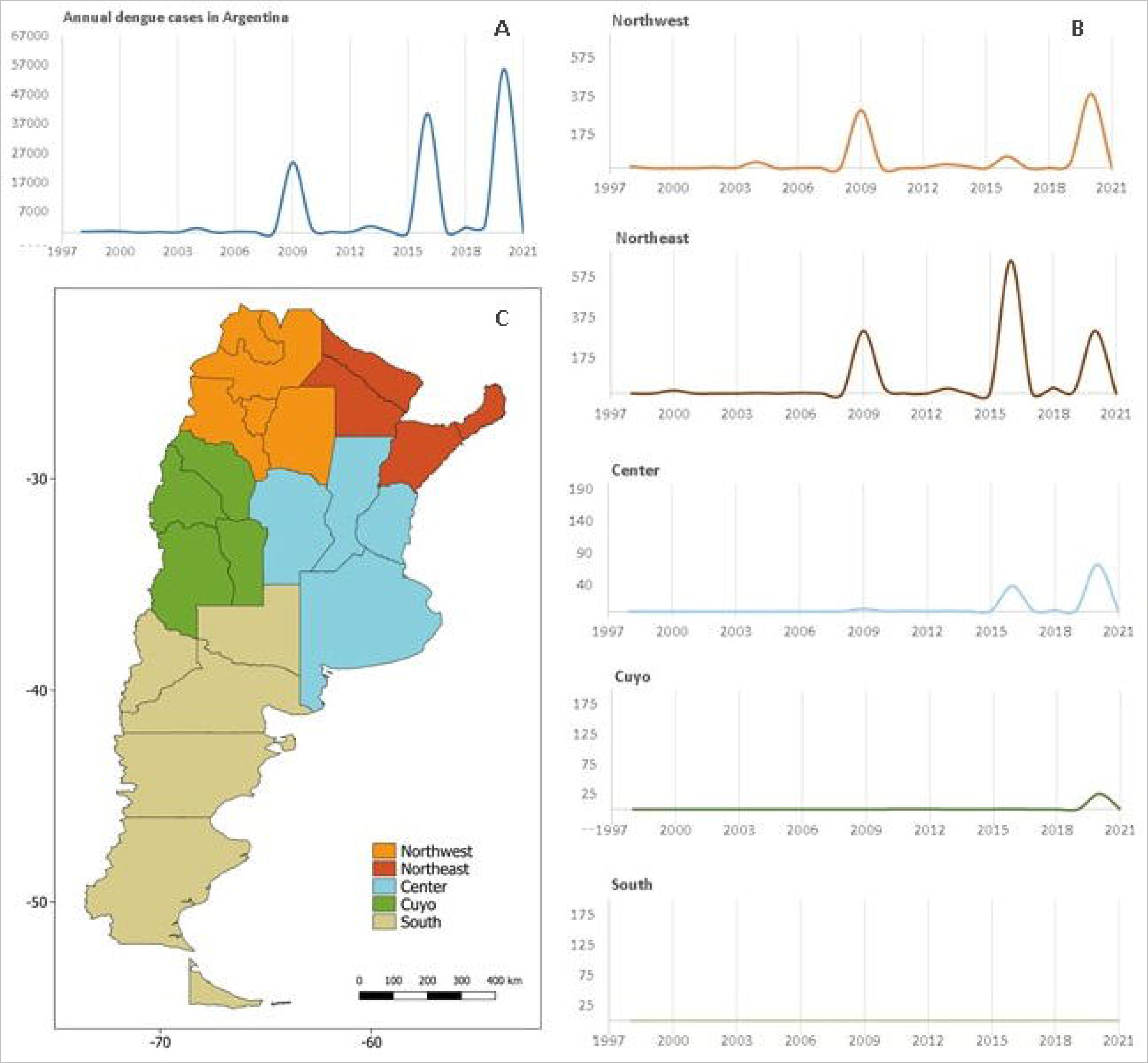
a) Cases of dengue in Argentina in the period 1998–2020 b) Incidence of DENV by geographic region (cases per 100,000 inhabitants) c) Geographic regions of Argentina

A time series was built with the number of cases per year to detect the epidemics that occurred in the study period. The incidence of DENV (number of cases per 100,000 inhabitants) was determined for each Argentinean region and maps were plotted to show the trends in registered epidemics. The incidence was also used to determine the most affected regions of Argentina. The maps were made with the QGis 3.14 software.

### Climate data

The climate variables analyzed were mean temperature, minimum temperature, maximum temperature, and precipitation from weather stations throughout the country. Data were provided by the National Meteorological Service (https://www.smn.gob.ar). Data were collected from 71 meteorological stations distributed in 20 provinces that presented autochthonous DENV cases. The climate indicators calculated for these variables were trends and anomalies, which were analyzed at the regional scale.

The climate trends were calculated as the slope of a linear regression of the precipitation and temperature time series during the 1961–2020 period. A positive (negative) slope means an average increase (decrease) of the mean values during that period. The trend maps were constructed with the results obtained for all the meteorological stations in the region.

The anomalies are defined as the difference between (mean, maximum, minimum) precipitation and temperature and the mean values of those variables in the reference period 1981–2010, established by the World Meteorological Organization (OMM, 2017). Mean anomalies were calculated for each province by averaging the anomaly values of each meteorological station in the province. A mean annual anomaly was then calculated using the mean values of all provinces with reported autochthonous DENV cases. To evaluate the changes in the climate variables, the anomalies of annual mean temperature, annual mean maximum temperature, and annual mean minimum temperature in two periods were compared; 1976–1997 (without DENV cases) and 1998–2020 (with DENV cases). These anomalies were analyzed using the Kruskal Wallis test (Infostat 2008).

According to Mordecai et al. (2019), the temperature range of DENV transmission is 17.8 – 34.5 °C. The number of months per year in 1976–2020 with that temperature range was counted at the meteorological stations closest to the provincial capitals and the autonomous city of Buenos Aires. Likewise, we analyzed the number of days per year with the optimal range of mean temperatures for transmission (28.4 – 29.8 °C, Mordecai et al. 2019). Subsequently, the means of the number of months and days were calculated for each meteorological station in the periods 1976–1997 (without DENV cases) and 1998–2020 (with DENV cases) to analyze changes in the number of months and days with optimal temperatures between periods.

## Results

Dengue cases in Argentina increased from the re-introduction of DENV in 1998 until the last and largest epidemic in 2020 (Figure 1a). During the 1998–2008 period, only five provinces reported autochthonous cases in the northeastern (NE) and northwestern (NW) country regions. In the period 2009–2020, the number of provinces with autochthonous cases rose to 20, adding the center, Cuyo, and south regions to the former regions identified. There were three large epidemics between 1998 and 2020, the first was in 2009 with 24,080 cases, the second in 2016 with 40,649 cases, and the last in 2020 with 55,854 cases. The NE and NW regions had the largest incidence in the period, with 1,344.14 and 790.92 cases (per 100,000 people), respectively. The central region had a total incidence of 123.33 cases (per 100,000 people), and Cuyo and southern Argentina had the lowest incidences with 27.27 and 0.3057 cases (per 100,000 people, Figure 1b), respectively. In four provinces (Río Negro, Chubut, Santa Cruz, and Tierra del Fuego) of the southern region, no DENV cases were registered. Figure 2 shows the increasing DENV incidence in the regions of Argentina since the reintroduction of the virus to the country until the last and most important epidemic of the year 2020.

**Figure 2.**
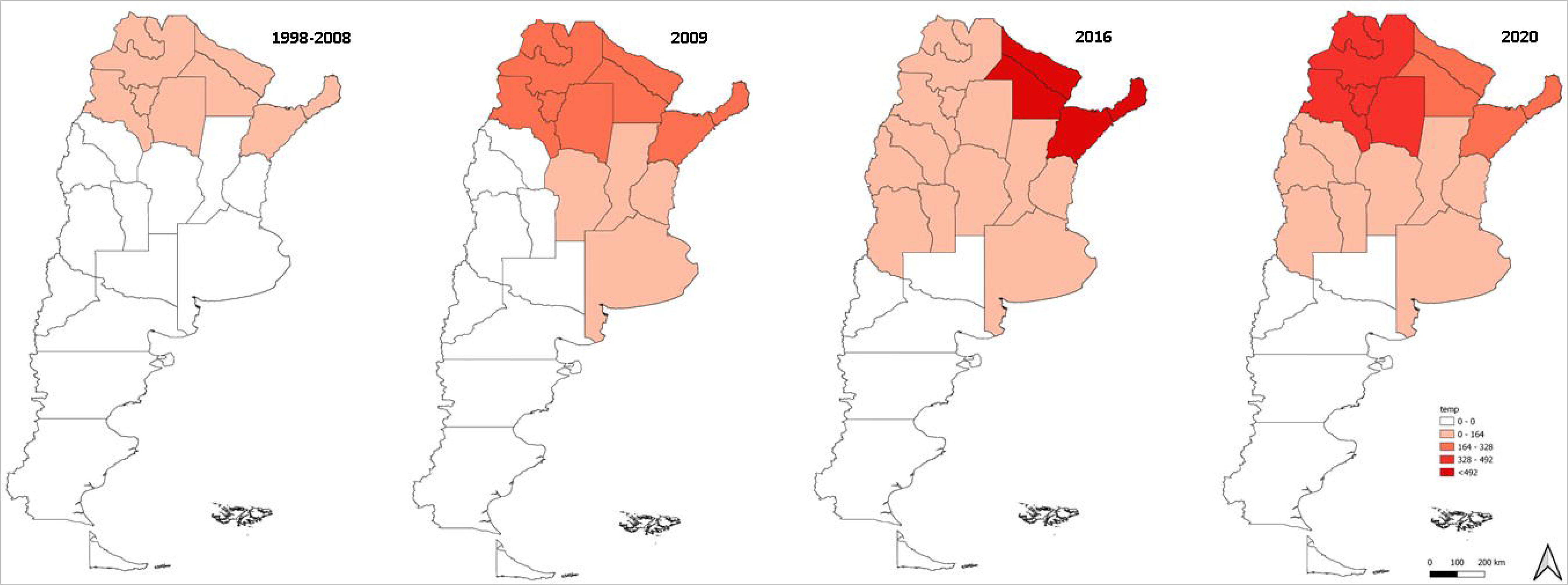
Incidence of DENV by region in the period 1998-2008 and in the three epidemics registered in 2009, 2016, and 2020.

The climate in Argentina underwent considerable changes in the past six decades. The annual mean, minimum, and maximum temperature trends show that there was a neutral (no increase) or positive variation (0– 1.5°C) in the country between 1961 and 2020 (Figure 3). The minimum temperature showed the greatest increase (1.5°C) over the geographic range compared to the rise in mean and maximum temperature (Figure 3). The precipitation trend showed a more erratic pattern, with decreases in northwestern, central-western, and southern provinces and increases in the central-eastern and northeastern provinces (Figure 3).

**Figure 3.**
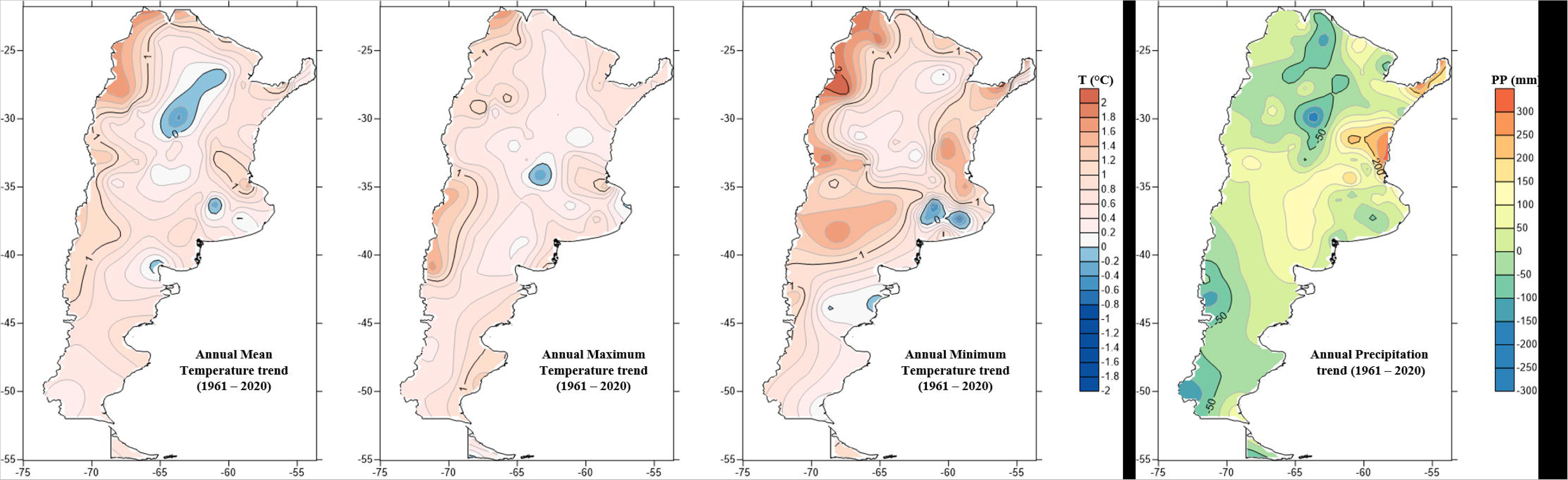
Trends of annual mean temperature, maximum temperature, minimum temperature and precipitation in the period 1961-2020 in Argentina.

When analyzing climate anomalies, temperature variables were significantly different between the periods under study (Table 1). No significant differences were observed in the precipitation of both periods (Table 1). The period of DENV transmission and epidemics (1998–2020) was characterized by warmer temperatures, with 82.60 % of the years (19 of 23) warmer mean temperatures, 73.91 % (17 of 23) warmer minimum, and 73.91 % (17 of 23) warmer maximum temperatures than the period without DENV transmission (1976–1997). Figure 4 shows the climate anomalies and autochthonous dengue transmission in the regions of Argentina from 1976 to 2020. The reintroduction of DENV into Argentina and its expansion to the central region coincide with the period of the greatest positive temperature anomalies in the country.

**Table 1.** Comparison of mean annual climatic anomalies in the periods 1976-1997 (without DENV cases, 22 years) and 1998-2020 (with DENV cases, 23 years). Asterisk (*) represents statistical significance at the α = 0.05. SD: standard deviation, H: statistical value.

| Variables | Period 1976-1997 |  | Period 1998-2020 |  | Statistic | Significance |
| --- | --- | --- | --- | --- | --- | --- |
|  | Mean | SD | Mean | SD |  |  |
| **Precipitation** | 1.45 | 11.28 | -1.63 | 11.94 | H = 1.50 | p = 0.2202 |
| **Mean temperature** | 0.08 | 0.38 | 0.43 | 0.38 | H = 7.86 | p = 0.0049* |
| **Minimum temperature** | -0.10 | 0.41 | 0.23 | 0.39 | H = 6.01 | p = 0.0139* |
| **Maximum temperature** | -0.09 | 0.44 | 0.33 | 0.55 | H = 6.35 | p = 0.0115* |

**Figure 4.**
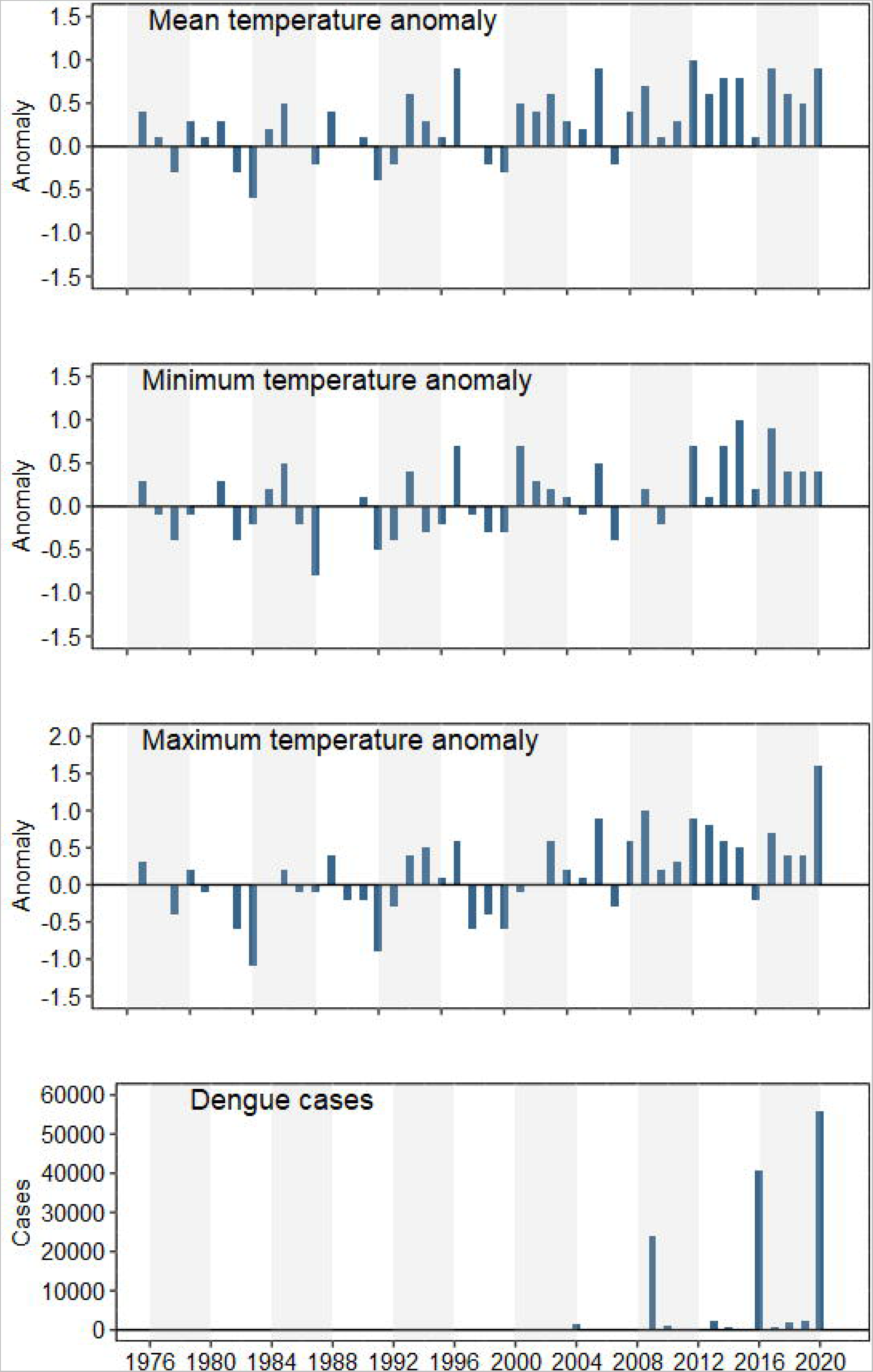
Temperature anomalies and autochthonous dengue cases in Argentina for the period 1976–2020.

The number of months with an optimal temperature range for the transmission of DENV was examined in the province’s capital cities in both periods (see Table 2). Some cities experienced a negative variation, e.g., Resistencia (in Chaco Province) with 1.34 fewer months with optimum transmission temperatures in the second period. The variation was positive in other cities, e.g., Neuquén, with an increase of 0.69. Although the number of months increased in some cities and decreased in others, the analysis of all the cities together reveals no positive or negative trend. Therefore, the results could indicate that the number of months with optimal temperatures for DENV transmission is constant in both periods in the analyzed cities.

**Table 2.**
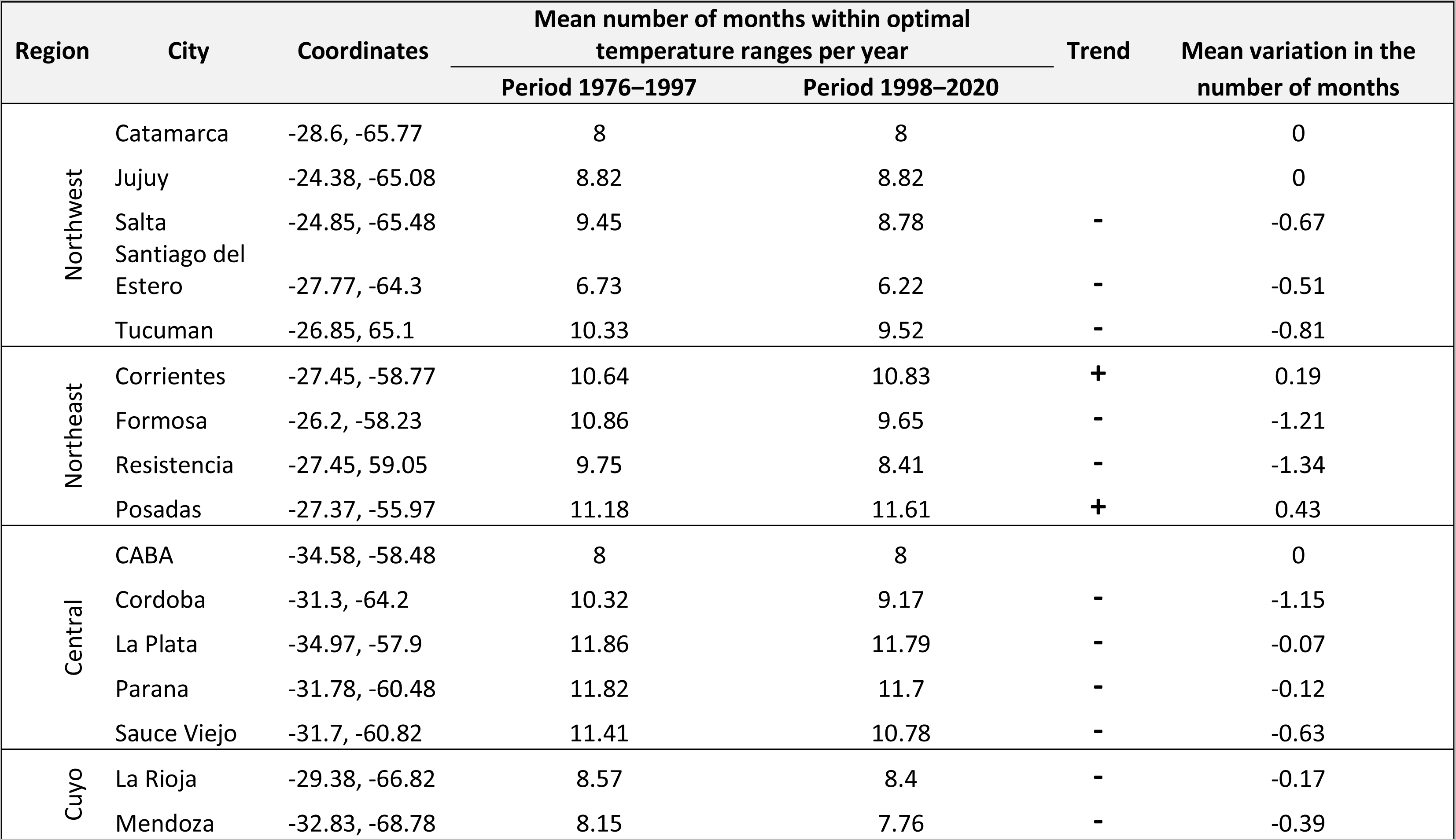
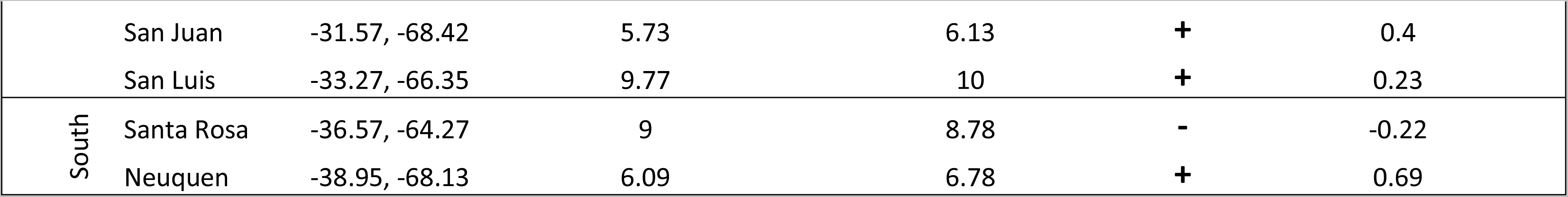
Difference in the mean number of months per year with optimal DENV transmission temperature ranges between periods 1976–1997 (without DENV cases) and 1998–2020 (with DENV cases). Trend: (+) indicates more months and (-) fewer months in 1998– 2020.

| Region | City | Coordinates | Mean number of months within optimal temperature ranges per year |  | Trend | Mean variation in the number of months |
| --- | --- | --- | --- | --- | --- | --- |
|  |  |  | Period 1976–1997 | Period 1998–2020 |  |  |
| Northwest | Catamarca | -28.6, -65.77 | 8 | 8 |  | 0 |
|  | Jujuy | -24.38, -65.08 | 8.82 | 8.82 |  | 0 |
|  | Salta | -24.85, -65.48 | 9.45 | 8.78 | - | -0.67 |
|  | Santiago del Estero | -27.77, -64.3 | 6.73 | 6.22 | - | -0.51 |
|  | Tucuman | -26.85, 65.1 | 10.33 | 9.52 | - | -0.81 |
| Northeast | Corrientes | -27.45, -58.77 | 10.64 | 10.83 | + | 0.19 |
|  | Formosa | -26.2, -58.23 | 10.86 | 9.65 | - | -1.21 |
|  | Resistencia | -27.45, 59.05 | 9.75 | 8.41 | - | -1.34 |
|  | Posadas | -27.37, -55.97 | 11.18 | 11.61 | + | 0.43 |
| Central | CABA | -34.58, -58.48 | 8 | 8 |  | 0 |
|  | Cordoba | -31.3, -64.2 | 10.32 | 9.17 | - | -1.15 |
|  | La Plata | -34.97, -57.9 | 11.86 | 11.79 | - | -0.07 |
|  | Parana | -31.78, -60.48 | 11.82 | 11.7 | - | -0.12 |
|  | Sauce Viejo | -31.7, -60.82 | 11.41 | 10.78 | - | -0.63 |
| Cuyo | La Rioja | -29.38, -66.82 | 8.57 | 8.4 | - | -0.17 |
|  | Mendoza | -32.83, -68.78 | 8.15 | 7.76 | - | -0.39 |
|  | San Juan | -31.57, -68.42 | 5.73 | 6.13 | + | 0.4 |
|  | San Luis | -33.27, -66.35 | 9.77 | 10 | + | 0.23 |
| South | Santa Rosa | -36.57, -64.27 | 9 | 8.78 | - | -0.22 |
|  | Neuquen | -38.95, -68.13 | 6.09 | 6.78 | + | 0.69 |

Table 3 shows the variation in the number of days with optimal temperatures for DENV transmission. An increase in the number of days within periods was observed, mainly in the cities of the central region where DENV cases increased in the past decade. In fact, an increased number of days was also seen in other cities like La Rioja (cuyo region), Posadas (northeast region), Resistencia (northeast region), and Salta (northwest region) which have had epidemics since the reintroduction of DENV in the country. DENV cases were also reported in cities in the southern region such as Santa Rosa and Neuquén in recentyears.

**Table 3.** Difference in the mean number of days per year with optimal DENV transmission temperature ranges between periods 1976– 1997 (without DENV cases) and 1998–2020 (with DENV cases). Trend: (+) indicates more days and (-) fewer days in 1998–2020.

| Region | City | Coordinates | Mean number of days with optimal temperatures |  | Trend | Mean variation in the<br>Period 1976-1997 |
| --- | --- | --- | --- | --- | --- | --- |
|  |  |  | Period 1976-1997 | Period 1998-2020 |  |  |
| Northwest | Catamarca | -28.6, -65.77 | 30.52 | 27.19 | - | 3.33 |
|  | Jujuy | -24.38, -65.08 | 37.32 | 33.64 | - | 3.68 |
|  | Salta | -24.85, -65.48 | 34.14 | 35.65 | + | 1.51 |
|  | Santiago del Estero | -27.77, -64.3 | 30.05 | 28.87 | - | 1.18 |
|  | Tucuman | -26.85, 65.1 | 30.16 | 29.96 | - | 0.2 |
| Northeast | Corrientes | -27.45, -58.77 | 36.68 | 35.05 | - | 1.63 |
|  | Formosa | -26.2, -58.23 | 35.19 | 33.35 | - | 1.84 |
|  | Resistencia | -27.45, 59.05 | 35.3 | 35.91 | + | 0.61 |
|  | Posadas | -27.37, -55.97 | 34.82 | 35.04 | + | 0.22 |
| Central | CABA | -34.58, -58.48 | 23.23 | 25.3 | + | 2.07 |
|  | Cordoba | -31.3, -64.2 | 28.86 | 31.55 | + | 2.69 |
|  | La Plata | -34.97, -57.9 | 20.95 | 22.27 | + | 1.32 |
|  | Parana | -31.78, -60.48 | 27.59 | 31.09 | + | 3.5 |
|  | Sauce Viejo | -31.7, -60.82 | 27.33 | 32.27 | + | 4.94 |
| Cuyo | La Rioja | -29.38, -66.82 | 27.76 | 32.1 | + | 4.35 |
|  | Mendoza | -32.83, -68.78 | 26.4 | 24.81 | - | 1.59 |
|  | San Juan | -31.57, -68.42 | 22.64 | 21.78 | - | 0.86 |
|  | San Luis | -33.27, -66.35 | 27.32 | 28.64 | + | 1.32 |
| South | Santa Rosa | -36.57, -64.27 | 21.05 | 21.39 | + | 0.34 |
|  | Neuquen | -38.95, -68.13 | 24.05 | 24.09 | + | 0.04 |

In summary, the number of months with optimal temperatures for DENV transmission was similar in the 1976–1997 and 1998–2020 periods in the analyzed cities. However, there was an increase in the number of days with an optimal temperature range that could be favoring the transmission in the cities analyzed between the 1976–1997 and 1998–2020 periods.

## Discussion

Central and South America are highly exposed, vulnerable, and strongly impacted by climate change (IPCC 2022). The impacts of climate and climate change on the dynamics, distribution, and spread of infectious diseases have received significant attention in recent years, as they can alter transmission seasonality and intensity (Huber et al. 2018; Robert et al. 2019; Rocklöv and Tozan 2019; DiSera et al. 2020; Robert et al. 2020). As a consequence of climate change, the incidence of climate-sensitive infectious diseases, particularly mosquito-borne diseases, is on the rise (Filho et al. 2019). This study shows how the temperature changes that occurred in Argentina during the past decades are associated with higher incidence and expansion of DENV in different regions of the country. This evaluation does not deny other factors involved in the expansion of DENV like greater connectivity and increased travel among countries, the rise in global population, or even improvement of public health systems (Stewart-Ibarra 2013; Barcellos and Lowe 2014; Rocklöv and Dubrow 2020). However, climate conditions allow the presence, abundance, and vector expansion, as well as higher virus transmission and the emergence of epidemics in areas where DENV is not endemic.

Studies were conducted on the optimal temperature ranges for the transmission of DENV by Ae. aegypti (Mordecai et al. 2019). Minimum temperatures were found to act as barriers, which impede DENV to spread into new regions (Butterworth et al. 2017; Lee et al. 2021). In addition, Díaz-Nieto et al. (2013) and Rubio et al. (2020) described the new distribution records for Ae. aegypti in southern south América. Our results show that the temperature trends of the past six decades are positive in most of the country, which is more remarkable for minimum temperatures, in agreement with other studies (Lovino et al. 2018a; Cogliati et al. 2021). This is consistent with an increase in the incidence and expansion of DENV in the different regions of Argentina (Estallo et al. 2014; Estallo et al. 2020; Robert et al. 2020; López et al. 2021). That is, the increase in temperature throughout the country has probably led to the circulation of the virus and the consequent increase in the frequency and magnitude of epidemics. Moreover, warmer temperatures may have triggered epidemics in areas without viral circulation until the past decade (Robert et al. 2019; Robert et al. 2020; Estallo et al. 2020; López et al. 2021). According to Messina et al. (2019), who examined DENV case occurrence related to climate, population, and socioeconomic projections, dengue risk is predicted to extend not only latitudinally but also to higher altitudes in northern Argentina between 2015 and 2080. Temperature (mean, minimum, and maximum) anomalies increased between the analyzed periods, with anomalies being higher in the period with DENV cases in Argentina. Monthly positive anomalies in minimum temperature have already been recorded in central Argentina, indicating that the period of dengue emergence in the city of Cordoba was characterized by warmer than average temperatures (Robert et al. 2020). These results show that the areas of Argentina prone to infectious diseases transmitted by mosquitoes could continue to expand and epidemics could consolidate in regions where outbreaks have already occurred.

The higher incidence of DENV in the different regions of Argentina does not show a relationship with precipitation trends and anomalies. This may be because the habits of people during droughts could be influencing the formation of mosquito breeding sites that generate epidemics, as mentioned in the literature (Estallo 2020; Stewart-Ibarra et al. 2020). For example, in 2020, the largest DENV epidemic was registered in Argentina while most of the country was under a strong precipitation deficit (negative anomalies) according to a report by the SMN (2020). In addition, the DENV epidemic coincided with the beginning of the COVID-19 pandemic and mandatory preventive social isolation. Consequently, vector control measuers were reduced and people remained in their homes exposed to Ae. aegypti much longer than usual, which according to Robert et al. (2020) could explain the greater magnitude of the DENV epidemic. Another explanation for the lack of relationship between precipitation and the increasing incidence of DENV in our results may be the regional scale of our analysis. The heavy precipitation episodes with which DENV outbreaks are associated (DiSera et al. 2020; Robert et al. 2020) generally occur over relatively small areas (local scale). In this sense, evaluating the cases based on precipitation on a smaller scale could show a better relationship between precipitation and the formation of outbreaks in the different regions of the country.

In the present work, cities across the country were evaluated to determine whether there had been an increase in the number of months and days with optimal mean temperatures for DENV transmission. Our results show no such increase in the number of months. However, the number of days with temperatures that contribute to maximizing transmission grew in most of the cities analyzed and in particular in the central region of the country. Given the important role that temperature plays in the timing of the extrinscic incubation period, having more days with optimal temperatures could be reducing the time taken by ingested viruses to develop to infective stages in mosquitoes, thereby increasing the spread rates of DENV (Sirisena and Noordeen 2014).

In agreement with other studies, we show that climate would increasingly favor the transmission of DENV, its expansion, and consolidation in new regions (Butterworth et al. 2017; Brady and Hay 2019; Estallo et al. 2020; López et al. 2021). In the past two decades, DENV and other viruses transmitted by Ae. aegypti mosquitoes rose dramatically and consistently with the combined effects of climate, urbanization, and declining vector control (Mordecai et al. 2019). The rapid warming of the Earth caused by anthropogenic greenhouse gas emissions has profound long-term implications for the prevention and control of vector-borne diseases (Rocklöv and Dubrow 2020). In this context, long-term monitoring and control programs are needed for Ae. aegypti and the detection of flaviviruses such as the one recently described by Cardozo et al. (2021) or the one proposed by Selvarajoo et al. (2022). These programs would make it possible to analyze the evolution of the vector and the circulation of flaviviruses. The early detection of arbovirus activity in new areas or increased viral activity in vector populations coupled with prompt medical attention is key to successfully controlling the associated diseases and reducing their case-fatality rate (Ribeiro da Silva et al. 2021). On the other hand, the simultaneous circulation of SARS-CoV-2 and arboviruses requires the integration of disease control measures and effective surveillance programs (Ribeiro da Silva et al. 2021). Climate warming may increase the geographic and seasonal ranges of mosquito-borne diseases with high thermal optima and upper limits relative to their current distribution. Current and future geographic range limits to transmission may depend primarily on the capacity of organisms to tolerate heat and cold stress, as well as factors like water availability, land use, and vector control (Mordecai 2019). Understanding the drivers of dengue expansion at the distribution boundaries is important for predicting whether dengue will continue to expand.

## Data Availability

All data produced in the present work are contained in the manuscript

## Acknowledgments

Funding for this study was provided through CAI+D 2020 (UNL) code 50520190100042LI. The data of dengue cases were granted by the National Ministry of Health, and meteorological data were granted by the National Meteorological Service of Argentina (SMN).

## References

Avilés G, Rangeón G, Vorndam V, Briones A, Baroni P, Enria D, et al. 1999. Dengue reemergence in Argentina. Emerg Infect Dis. 5, 575–8.

Brathwaite Dick O, San Martín JL, Montoya RH, del Diego J, Zambrano B, Dayan GH. 2012. The history of dengue outbreaks in the Americas. Am. J. Trop. Med. Hyg. 87, 584–93.

Bhatt S, Gething PW, Brady OJ, Messina JP, Farlow AW, Moyes CL, et al. 2013. The global distribution and burden of dengue. Nature 496, 504–507.

Boggs, C. L. & Inouye, D. W. A single climate driver has direct and indirect effects on insect population dynamics. Ecol. Lett. 15, 502–508 (2012).

Brady, O. J. & Hay, S. I. The global expansion of dengue: How Aedes aegypti mosquitoes enabled the first pandemic arbovirus. Annu. Rev. Entomol. 65, 191–208 (2020).

Butterworth, M. K., Morin, C. W. & Comrie, A. C. An analysis of the potential impact of climate change on dengue transmission in the southeastern United States. Environ. Health. Perspect. 125, 579–585 (2017).

Caldwell, J. M. et al. Climate predicts geographic and temporal variation in mosquito-borne disease dynamics on two continents. Nature Comm. https://doi.org/10.1038/s41467-021-21496-7 (2021).

Cardozo, F. et al. Implementación de un sistema de detección de flavivirus en mosquitos. Mem. Inst. Investig. Cienc. Salud. 19, 32–40 (2021).

Carrington, L. B., Armijos, M. V., Lambrechts, L. & Scott, T. W. Fluctuations at a low mean temperature accelerate dengue virus transmission by Aedes aegypti. PLoS Negl. Trop. Dis. 7 e2190 (2013).

Cogliati, M., Müller, G. V. & Lovino, M. A. Seasonal trend analysis of minimum air temperature in La Plata River Basin. Theo. and App. Clim. 144, 25–37 (2021).

Díaz-Nieto, L. M., Maciá, A., Perotti, M. A. & Berón, C. M. Geographical limits of the southeastern distribution of Aedes aegypti (Diptera, Culicidae) in Argentina Leonardo. PLoS Neg. Trop. Dis. 7, e1963 (2013).

DiSera, L. et al. The Mosquito, the virus, the climate: an unforeseen réunion in 2018. GeoHealth. 4, e2020GH000253. (2020).

Estallo, E. L. Factores eco-epidemiológicos asociados a la distribución y abundancia de mosquitos vectores de arbovirus. In Arbovirosis de importancia en las regiones tropicales, Vol. 5, pp:154–172. (CIDEPRO, Ecuador, 2020).

Ferreira, G. L. C. Global dengue epidemiology trends. Rev. Inst. Med. Trop. São Paulo. 54, 5–6 (2012).

Gubler, D. J. The global emergence/resurgence of arboviral diseases as public health problems. Arch Med Res. 33, 330–42 (2002).

Gubler, D. J. Dengue, urbanization and globalization: the unholy trinity of the 21st century. Trop. Med. Heal. 39, 3–11 (2011).

Hii, Y.L., Rocklöv, J., Ng, N., Tang, C. S., Pang, F. Y. & Sauerborn R. Climate variability and increase in intensity and magnitude of dengue incidence in Singapore. Glob. Heal. Act. https://doi.org/10.3402/gha.v2i0.2036 (2009).

Huber, J.H., Childs, M. L., Caldwell, J. M. & Mordecai, E. A. Seasonal temperature variation influences climate suitability for dengue, chikungunya, and Zika transmission. PLoS Negl. Trop. Dis. 12, e0006451 (2018).

InfoStat. InfoStat, versión 2008. Manual del Usuario. Grupo InfoStat, FCA, Universidad Nacional de Córdoba. Primera Edición, Editorial Brujas Argentina. (2008).

IPCC. Summary for Policymakers. Climate Change 2021: The Physical Science Basis. Preprint at https://www.ipcc.ch/report/sixth-assessment-report-working-group-i/ (2021).

IPCC. Summary for Policymakers. Climate Change 2022: Impacts, adaptation and vulnerability. Preprint at https://www.ipcc.ch/report/ar6/wg2/ (2022).

Kraemer, M. U. G. et al. The global distribution of the arbovirus vectors Aedes aegypti and Ae. albopictus. eLife. 4:e08347 (2015).

Lambrechts, L., Scott, T. W. & Gubler, D. J. Consequences of the expanding global distribution of Aedes albopictus for dengue virus transmission. PLoS. Negl. Trop. Dis. 25; e646 (2010).

La Ruche, G. et al. First two autochthonous dengue virus infections in metropolitan France, September 2010. Euro Surveill. 15, http://www.eurosurveillance.org/ViewArticle.aspx?ArticleId=19676 (2010).

Lee, S. A. EconomouI, T., Castro Catão, R., Barcellos, C. & Lowe R. The impact of climate suitability, urbanisation, and connectivity on the expansion of dengue in 21st century Brazil. PLoS Negl. Trop. Dis. https://doi.org/10.1371/journal.pntd.0009773 (2021).

López, M. S. et al. Dengue arbovirus affecting temperate Argentina province for more than a decade 2009-2020. Sc. Data. https://doi.org/10.1038/s41597-021-00914-x (2021).

Lovino, M. A., Müller, O., Berbery, E. H. & Müller, G. How have daily climate extremes changed in the recent past over northeastern Argentina? Glob. Planet. Chang. 168, 78–97. https://doi.org/10.1016/j.gloplacha. (2018a)

Lovino, M. A., Müller, O. V., Müller, G. V., Sgroi, L. C. & Baethgen, W. E. Interannual-to-multidecadal hydroclimate variability and its sectoral impacts in northeastern Argentina. Hydrol. Earth. Syst. Sci. 22, 3155–3174 (2018b).

Messina, J. P. et al. The current and future global distribution and population at risk of dengue. Nature Micr. 4, 1508–1515 (2019).

MHO (Ministry Health Organization). Directrices para la prevención y control de Aedes aegypti. Preprint at https://bancos.salud.gob.ar/recurso/directrices-para-la-prevencion-y-control-de-aedes-aegypti (2016).

MHO (Ministry Health Organization). Dengue - Algoritmo de diagnóstico y notificación. Preprint at https://bancos.salud.gob.ar/recurso/dengue-algoritmo-de-diagnostico-y-notificacion (2016).

Mordecai, E. A. et al. Thermal biology of mosquito-borne disease. Ecol. Lett. 22, 1690–1708 (2019).

Murray, N. E. A., Quam, M. B. & Wilder-Smith, A. Epidemiology of dengue: past, present and future prospects. Clin. Epidemiol. 20, 299–309 (2013).

WHO (World Health Organization). Dengue and severe dengue. Preprint at https://www.who.int/news-room/fact-sheets/detail/dengue-and-severe-dengue

Messina, J. P. et al. Global spread of dengue virus types: mapping the 70 year history. Trends Microbiol. 22, 138–146 (2014).

Patterson, J., Sammon, M. & Garg, M. Dengue, Zika and chikungunya: emerging arboviruses in the New World. West J. Emerg. Med. 17, 671–9 (2016).

Rey, J. R. Dengue in Florida (USA). Insects. 5, 991–1000 (2014).

Ribeiro da Silva, S. J., Ferraz de Magalhaes, J. & Pena, L. Simultaneous Circulation of DENV, CHIKV, ZIKV and SARS-CoV-2 in Brazil: an Inconvenient Truth. One Health 12, 100205 (2021).

Robert, M.A. et al. Arbovirus emergence in the temperate city of Córdoba, Argentina, 2009–2018. Sci. Data. 21, https://doi.org/10.1038/s41597-019-0295-z (2019).

Robert, M. A., Stewart-Ibarra, A. M. & Estallo, E. L. Climate change and viral emergence: evidence from Aedes-borne arboviruses. Curr. Op. in Vi. 40, 1–7 (2020).

Rocklöv, J. & Tozan, Y. Climate change and the rising infectiousness of dengue. Em. Top. Life Sci. doi:10.1042/etls20180123 (2019).

Rocklöv, J. & Dubrow, R. Climate change: an enduring challenge for vector-borne disease prevention and control. Nature Imm. 21, 479–483 (2020)

Rubio, A., Cardo, M. V., Vezzani, D. & Carbajo, A. E. Aedes aegypti spreading in South America: new coldest and southernmost records. Mem. Inst. Osw. Cruz. 115, e190496 (2020).

Selvarajoo, S. et al. Dengue surveillance using gravid oviposition sticky (GOS) trap and dengue non-structural 1 (NS1) antigen test in Malaysia: randomized controlled trial. Sc. Rep. 12, https://doi.org/10.1038/s41598-021-04643-4 (2022).

SMN (National Metereological Service). Monitoreoo de la precipitación en Argentina año 2020. Preprint at https://www.smn.gob.ar/sites/default/files/monitoreo_precipitaci%C3%B3n_a%C3%B1o2020.pdf (2020)

Stewart-Ibarra, A. M., Romer, M. & Borbor-Cordova, M. J. El rol de los sistemas socioecológicos en la transmisión de arbovirus: caso estudio en Ecuador. In Arbovirosis de importancia en las regiones tropicales, Vol. 6, pp:181–211. (CIDEPRO, Ecuador, 2020).

PHCC. National Action Plan for Health and Climate Change 2019. Preprint at http://servicios.infoleg.gob.ar/infolegInternet/anexos/330000-334999/332234/res447-6.pdf (2019).

Sirisena, P. D. N. N. & Noordeen, F. Evolution of dengue in Sri Lanka—changes in the virus, vector, and climate. Int. J. of Inf. Dis. 19, 6–12 (2014).

Tomasello, D. & Schlagenhauf, P. Chikungunya and dengue autochthonous cases in Europe, 2007–2012. Tra. Med. Infect. Dis. 11, 274–84 (2013).

Vezzani, D. & Carbajo, A. E. Aedes aegypti, Aedes albopictus, and dengue in Argentina: current knowledge and future directions. Mem Inst Osw. Cruz. 31, 66–74 (2008).

Wilder-Smith, A. & Gubler, D. J. Geographic Expansion of Dengue: The Impact of International Travel. Med. Clin. North Am. 92, 1377–1390 (2008).

WHO (World Health Organization). Dengue y dengue grave. Preprint at https://www.who.int/news-room/fact-sheets/detail/dengue-and-severe-dengue.

WMO. (World Meteorological Organization). Directrices de la Organización Meteorológica Mundial sobre el cálculo de las normales climáticas (WMO-No. 1203). Ginebra. ISBN 978-92-63-311203-7 (2017).

